# Maternal micronutrient deficiencies and inflammation and their associations with adverse birth outcomes: The BRINDA project

**DOI:** 10.64898/2026.05.26.26353988

**Authors:** Jiaxi Geng, Hanqi Luo, Rochelle Werner, Lei Liu, Yaw Addo, Usha Ramakrishnan, Maria J. Ramirez-Luzuriaga, Phuong Hong Nguyen, Parminder S Suchdev, Melissa F Young, Yi-An Ko

## Abstract

**Background:** Maternal micronutrient deficiencies (MNDs) and inflammation contribute to adverse birth outcomes While the individual effects of MNDs have been studied, the consequence of co-occurring MNDs remains unclear.

**Objectives:** To examine the associations between maternal micronutrient deficiencies and inflammation with adverse birth outcomes (ABOs).

**Methods:** Data from 5,408 pregnant women across 11 datasets from 10 countries were analyzed. Descriptive analyses explored the distribution of MNDs (iron, vitamin A, zinc, serum folate, vitamin D, and vitamin B12) and inflammation (c-reactive protein >5 mg/L or α-(1)-acid glycoprotein > 1g/L) by maternal characteristics (age, height, education, socioeconomic status [SES]) using chi-square tests. Associations of 1) single MNDs and inflammation and 2) co-occurring MNDs (2 deficiencies at a time) with low birth weight (LBW, < 2500 g), preterm birth (PTB, < 37 wks), and small-for-gestational age (SGA, < 10th percentile for gestational age), were examined using modified Poisson regression to estimate relative risk (RR), adjusting for age, SES, and dataset.

**Results:** Young maternal age and short height were associated with up to 9.7% and 25% higher prevalence of MNDs and inflammation, respectively. Lower education and SES level were associated with higher prevalence of Vitamin B12 deficiency. Women with folate deficiency had an increased risk of LBW (RR [95% CI]: 1.22 [1.06, 1.39]). Co-occurring MNDs for folate and vitamin B12 were also associated with increased LBW risk (1.38 [1,1.9]) as was folate deficiency without iron (1.28 [1.09, 1.51]) or vitamin B12 deficiency (1.67 [1.09, 2.56]) compared with mothers without either deficiency. Iron deficiency without vitamin B12 deficiency was associated with a reduced LBW risk (0.4 [0.2, 0.79]).

**Conclusion:** Maternal MNDs, especially folate and vitamin B12, are linked to adverse birth outcomes. Complex nutrient interactions highlight the need to explore these relationships to improve maternal and neonatal health interventions.

## Introduction

Adverse birth outcomes, including low birth weight (LBW), preterm birth (PTB), and being small for gestational age (SGA), are significant global health challenges that impact neonatal health^1^ and are associated with increased neonatal mortality, morbidity, and long-term developmental challenges for the child^2^.

Understanding the role of maternal nutritional status is crucial for unraveling potential links to adverse birth outcomes.^3^ Several studies have explored the associations between individual micronutrient deficiencies (MNDs) and birth outcomes among pregnant women. For example, iron deficiency is one of the most prevalent MNDs in pregnancy^4^, and iron supplementation during pregnancy has been associated with increased birth weight^5^. Zinc supplementation has been associated with a modest 14% reduction in PTB risk, although it showed limited impact on LBW or SGA^5^. Inadequate folate levels have also been linked to increased risks of PTB and LBW^6^.

Despite growing evidence, most prior research has focused on the impact of individual micronutrients in isolation; however, many pregnant women, particularly in low-resource settings, experience multiple concurrent MNDs. The combined effects of these deficiencies on birth outcomes remain underexplored. A recent review concluded that supplementation with multiple micronutrients (MMN), including iron and folic acid, substantially reduced the risk of LBW and slightly decreased the risk of PTB and SGA.^7^ While supplementation trials have compared MMN versus iron–folic acid interventions^8^, limited studies have directly examined how maternal biomarker-based MMN status relates to specific birth outcomes and the role inflammation plays in those interactions.

The present study addresses this gap by using data from the BRINDA (Biomarkers Reflecting Inflammation and Nutritional Determinants of Anemia) project to: 1) quantify the individual effects of selected micronutrients (iron, vitamin A, zinc, serum folate, vitamin D, and vitamin B12) and inflammation on ABOs, and 2) assess the combined effects of multiple MNDs on LBW, PTB, and SGA. Findings from the study can inform the development of maternal nutrition strategies and policies aimed at improving maternal health and neonatal outcomes.

## Methods

### Data Source

The Biomarkers Reflecting Inflammation and Nutritional Determinants of Anemia (BRINDA) Project (https://www.brinda-nutrition.org/) aims to improve global assessment of micronutrient status. **Supplementary Figure 1** shows the stepwise inclusion criteria used to derive the final study sample of pregnant women. The BRINDA project harmonized data from 26,632 pregnant women across 20 datasets. Of these, 12,189 women had birth outcome data. This analysis included 5,408 women from 11 datasets who had at least one measurement of inflammation and one micronutrient (iron, vitamin A, zinc, serum folate, vitamin B12, or vitamin D) and attended at least two visits during pregnancy. These datasets originated from 10 countries: Bangladesh^9,8^, the Gambia^10^, Ghana^11^, Malawi^12^, Mexico^13^, the United States^14^, Vietnam^15^, Guatemala^16^, India^16^, and Pakistan^16^. This repeated measures database included women had two (n=5097) or three (n=311) visits throughout pregnancy, resulting in a total of 11,127 observations. All datasets, except for one cohort study, were derived from randomized controlled trials in which women received supplementation with folic acid, iron and folic acid combinations, lipid-based nutrients, or multiple micronutrients including calcium, potassium, magnesium, and essential fatty acids.

### Birth Outcomes

Gestational age at delivery and birth weight were recorded at delivery. Gestational age was determined based on the time interval between the mother’s last menstrual period and the child’s date of birth. LBW was defined as a birth weight less than 2500 grams, PTB was defined as a gestational age at delivery less than 37 weeks, and SGA was defined as a birth weight less than the 10th percentile for gestational age in days according to the 21st Intergrowth guidance^17^.

### MNDs and Inflammation

Inflammation was defined as the elevated C-reactive protein [CRP, > 5 mg/L] or α-1-acid glycoprotein [AGP, > 1 g/L]. Iron deficiency was defined by inflammation-adjusted serum ferritin (SF) levels < 15 µg/L or soluble transferrin receptor (sTfR) levels > 8.3 mg/L^18^. Vitamin A deficiency was defined as serum retinol (SR) or retinol binding protein (RBP) levels < 0.7 µmol/L^19^. Zinc deficiency was defined based on the timing and fasting status of the blood draw: < 700 µg/L for fasting morning draws, < 660 µg/L for non-fasting morning draws, or < 590 µg/L for afternoon or unspecified times in case fasting status was unknown^20^. Serum folate (SFO) deficiency was defined as serum folate levels < 10 nmol/L^21^. Vitamin D deficiency was defined as 25-hydroxyvitamin D (25(OH)D) levels < 25 nmol/L^22^. Vitamin B12 deficiency was defined as vitamin B12 levels < 220 pmol/L^23^.

### Laboratory Methods

Biomarkers were measured from venous or capillary blood samples. Venous samples were centrifuged, and plasma or serum aliquots were stored at temperatures below −20°C until analysis. The measurement methods differed by biomarker and dataset have been described in the online supplementary materials.^24^

### Statistical Analysis

Maternal characteristics were summarized by dataset using quartiles and percentages. SES was generated from the asset index developed using principal components analysis (PCA) or predefined quintiles, with values from 0 to 4. SES was then categorized into three levels: low, medium, and high based on asset quintiles. We compared the prevalence of each MND and the prevalence of inflammation at baseline visit across different demographic risk factors, including age, height, education, SES, and smoking status using chi-square test.

To assess the individual effects of MNDs and inflammation on birth outcomes, we classified women as “exposed” if they had measurement indicating MND or inflammation at any time point during pregnancy. This approach captures whether exposure at any time point during pregnancy is associated with outcomes. To examine exposures later in pregnancy, the analysis was repeated using biomarker data from the last available visit (n=5408), which occurred in the second or third trimester (median gestational age: 39 weeks; range: 24–43 weeks). Sample sizes for each micronutrient are provided in **Supplementary Figure 1**. Modified Poisson regression^25^ with robust standard error was used to estimate adjusted relative risks (RRs) for each adverse birth outcome, controlling for maternal age, SES, and dataset.

To examine the combined effects of co-occurring deficiencies, we restricted the analysis to women with data on at least two different micronutrient biomarkers. To minimize bias and overfitting, we only analyzed biomarker pairs that met the following a priori criteria: 1) available from at least two datasets; 2) ≥30 women in each exposure category (no deficiency, one deficiency, and two deficiencies); and 3) ≥20 women with the specific birth outcome. These criteria resulted in 5 unique combinations.

All analyses were conducted using R version 4.3.1. False discovery rate was used to account for multiple tests^26^. An adjusted p-value of ≤ 0.05 was considered statistically significant.

## Results

### Descriptive statistics

**Table 1** shows maternal characteristics across 11 datasets. Demographic characteristics varied widely among included datasets, from a median of 19 years to 29 years for maternal age and 1.2% (the Gambia) to 82.9% (Guatemala) for the percentage of women with height ≤150 cm. Percentage of three adverse birth outcomes varied as LBW (5% to 42.8%), PTB (1.3% to 27%) and SGA (7.7% to 63.8%). In terms of inflammation and MNDs, the percentage of women with inflammation ranged from 6.9% to 73.8%. Iron deficiency ranged from 1.5% to 90.3%. Vitamin A deficiency was 1.8% and 7% in the two available datasets. Zinc deficiency ranged from 13.5% to 82.7%, folate deficiency from 1.8% to 12.5%, vitamin B12 deficiency from 3.7% to 68.7%. Vitamin D deficiency was 1.4% and 8.5% in the two available datasets.

**Table 1:** Baseline characteristics, prevalence of adverse birth outcome, inflammation, and micronutrient deficiencies of pregnant women by dataset.

|  | Bangladesh(2012) | Bangladesh(2013) | Ghana(2014) | The Gambia (2013) | Guatemala(2018) | India(2017) | Malawi(2012) | Mexico(2003) | Pakistan(2017) | US(2013) | Vietnam(2011) |
| --- | --- | --- | --- | --- | --- | --- | --- | --- | --- | --- | --- |
| n | 1291 | 2808 | 995 | 689 | 556 | 554 | 1042 | 606 | 627 | 233 | 1311 |
| Age, y [median(Q1,Q3)] | 21(18, 25) | 21(18, 25) | 26(23, 30) | 29(24, 34) | 24(21, 27) | 21(19, 24) | 24(20, 29) | 22(19, 26) | 24(20, 27) | 19(17, 28) | 26(23, 29) |
| Rural or Urban | Urban | Rural | Rural | Semiurban | - | - | Rural | Semiurban | - | Urban | Rural |
| Height <= 150cm, n(%) | 750(58.1%) | 1234(43.9%) | 51(5.1%) | 8(1.2%) | 461(82.9%) | 226(40.8%) | 131(12.6%) | 368(60.7%) | 212(33.8%) | 7(3%) | 391(29.8%) |
| **Socioeconomic status, n(%)** |  |  |  |  |  |  |  |  |  |  |  |
| Low | 515(39.9%) | 1067(38%) | 489(49.1%) | - | 62(11.2%) | 54(9.7%) | 411(39.4%) | 208(34.3%) | 303(48.3%) | - | 509(38.8%) |
| Medium | 523(40.5%) | 1140(40.6%) | 298(29.9%) | - | 332(59.7%) | 360(65%) | 426(40.9%) | 193(31.8%) | 228(36.4%) | - | 548(41.8%) |
| High | 253(19.6%) | 601(21.4%) | 207(20.8%) | - | 162(29.1%) | 140(25.3%) | 185(17.8%) | 114(18.8%) | 96(15.3%) | - | 254(19.4%) |
| **Adverse birth outcome, n(%)** |  |  |  |  |  |  |  |  |  |  |  |
| Low Birth Weight | 552(42.8%) | 1033(36.8%) | 81(8.1%) | 66(9.6%) | 75(13.5%) | 145(26.2%) | 109(10.5%) | 56(9.2%) | 195(31.1%) | 62(26.6%) | 66(5%) |
| Preterm | 188(14.6%) | 264(9.4%) | 21(2.1%) | 9(1.3%) | 35(6.3%) | 25(4.5%) | 26(2.5%) | 58(9.6%) | 99(15.8%) | 63(27%) | 42(3.2%) |
| Small for Gestational Age | 770(59.6%) | 1792(63.8%) | 229(23%) | 193(28%) | 131(23.6%) | 269(48.6%) | 249(23.9%) | 142(23.4%) | 230(36.7%) | 18(7.7%) | 197(15%) |
| **Micronutrient deficiency or inflammation, n(%)** |  |  |  |  |  |  |  |  |  |  |  |
| Inflammation | 202(16.1%) | 116(16.3%) | 375(38.3%) | 73(23.2%) | 149(73.8%) | 81(44.8%) | 438(42.6%) | 104(27.3%) | 65(34%) | 77(42.8%) | 86(6.9%) |
| Iron Deficiency | 106(8.5%) | 90(12.6%) | 131(13.4%) | 147(45.7%) | 6(3%) | 90(58.1%) | 279(27.1%) | 344(90.3%) | - | 102(46.2%) | 18(1.5%) |
| Vitamin A Deficiency | 86(7%) | - | - | - | - | - | - | - | - | - | 22(1.8%) |
| Zinc Deficiency | 167(13.5%) | - | - | - | 117(58.2%) | 102(56.4%) | - | - | 158(82.7%) | - | - |
| Folate Deficiency | 157(12.5%) | - | 32(10.9%) | - | - | - | - | - | - | 3(1.8%) | - |
| Vitamin B12 Deficiency | 837(68.7%) | - | 11(3.7%) | - | - | - | - | - | - | 30(21.9%) | - |
| Vitamin D Deficiency | 17(1.4%) | - | - | - | - | - | - | - | - | 12(8.5%) | - |

1. Q1, 25% percentile; Q3, 75% percentile. 2. Low birth weight was defined as a birth weight of < 2500 grams; Preterm birth was defined as a gestational age at delivery < 37 weeks; and small-for-gestational age was defined as a birth weight < the 10th percentile for gestational age according to the 21st Intergrowth guidance (http://intergrowth21.ndog.ox.ac.uk/). 3. Inflammation was defined as C-reactive protein (CRP) > 5 mg/L, or α-1-acid glycoprotein (AGP) > 1 g/L; iron deficiency was defined by inflammation-adjusted serum ferritin levels < 15 μg/L or transferrin receptor levels > 8.3 mg/L; vitamin A deficiency was defined as retinol binding protein or serum retinol levels < 0.7 μmol/L; zinc deficiency was defined based on the timing and fasting status of the blood draw, along with the zinc concentration: < 700 μg/L for fasting morning draws, < 660 μg/L for non-fasting morning draws or if fasting status was unknown, and < 590 μg/L for afternoon or unspecified times; serum folate deficiency was defined as serum folate levels < 10 nmol/L; vitamin D deficiency was defined as 25-hydroxyvitamin D (25(OH)D) levels < 25 nmol/L; vitamin B12 deficiency was defined as vitamin B12 levels < 20 pmol/L.

The prevalence of MNDs and inflammation varies across different demographic and risk factors. **Supplementary Figure 2** displays stratified results by dataset. Women aged ≤ 20 years showed a significantly higher prevalence of iron (21.9% vs. 18.7%), vitamin A (6% vs. 3.7%), vitamin B12 (58.5% vs. 48.8%), and vitamin D (2.9% vs. 1.1%) deficiencies, but lower prevalence of zinc (24.9% vs. 34%) and folate (8.9% vs. 13.1%) deficiencies compared to older women (each p<0.05). Women of shorter height (≤ 150 cm) had significant higher prevalence of iron (23.2% vs. 18.4%, p<0.05), vitamin A (7.5% vs. 3.1%, p<0.05), and vitamin B12 (67.5% vs. 41.9%, p<0.05) deficiencies. Higher education was associated with lower prevalence of inflammation, vitamin A, and vitamin B12 deficiencies. SES appeared to be associated with vitamin B12 deficiency, with the highest prevalence observed in the middle SES group (60.2%) and the lowest in the high SES group (47.7%).

### Individual MND and Adverse Birth Outcomes

**Figure 1** shows the percentages of three adverse birth outcomes by MND and inflammation status. Results are based on inflammation or deficiency at any point during pregnancy. LBW was significantly more common among women deficient in vitamin A (45.7% vs. 22%, p<0.001), folate (43.2% vs.32.3%, p<0.001), and vitamin B12 (40.7% vs. 21.2%, p<0.001), while deficiency in zinc was related to significantly lower prevalence of LBW (30.8% vs. 39.7%, p<0.001). Increased PTB was significantly associated with iron deficiency (7.62% vs. 6.17%, p=0.025). SGA prevalence higher among women with vitamin A (61.3% vs. 35.4%, p<0.001), folate (54.5% vs. 45.2%, p=0.002), vitamin B12 (55.9% vs. 29.6%, p<0.001) deficiencies, but lower with inflammation (31.3% vs. 34.7%, p=0.0042) and zinc deficiency (44.7% vs. 58.1%, p<0.001). Results using data from the last study visit were consistent with earlier findings (**Supplementary Figure 3)**. There was a significantly higher prevalence of PTB among women with inflammation (8.15% vs. 6.27%, p=0.016) and vitamin A deficiency (13.6% vs. 8.54%, p=0.024). Lower prevalence of SGA was observed with maternal inflammation (31.2% vs. 40.6%, p<0.001), iron deficiency (35.3% vs. 39.2%, p=0.005), and vitamin D deficiency (36.5% vs. 55.6%, p=0.01).

**Figure 1:**
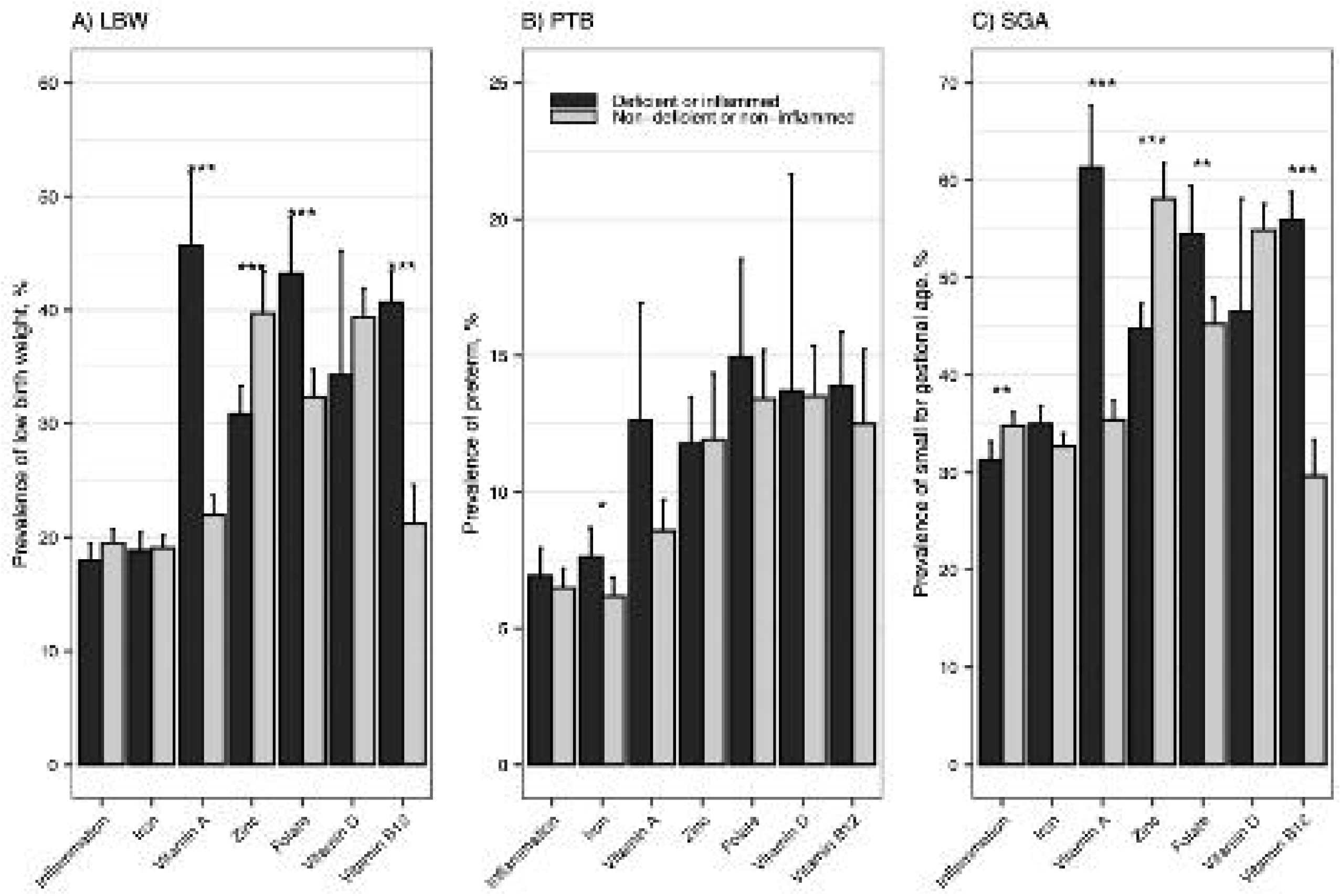
Percentages of (A) LBW, (B) PTB, and (C) SGA by inflammation and micronutrient status at any time point during pregnancy 1. LBW, low birth weight; PTB, preterm birth; SGA, small for gestational age. 2. LBW was defined as a birth weight of < 2500 grams; PTB was defined as a gestational age at delivery < 37 weeks; and SGA was defined as a birth weight < the 10th percentile for gestational age according to the 21st Intergrowth guidance (http://intergrowth21.ndog.ox.ac.uk/). 3. Inflammation was defined as C-reactive protein (CRP) > 5 mg/L, or α-1-acid glycoprotein (AGP) > 1 g/L; iron deficiency was defined by inflammation-adjusted serum ferritin levels < 15 µg/L or transferrin receptor levels > 8.3 mg/L; vitamin A deficiency was defined as retinol binding protein or serum retinol levels < 0.7 µmol/L; zinc deficiency was defined based on the timing and fasting status of the blood draw, along with the zinc concentration: < 700 µg/L for fasting morning draws, < 660 µg/L for non-fasting morning draws or if fasting status was unknown, and < 590 µg/L for afternoon or unspecified times; serum folate deficiency was defined as serum folate levels < 10 nmol/L; vitamin D deficiency was defined as 25-hydroxyvitamin D (25(OH)D) levels < 25 nmol/L; vitamin B12 deficiency was defined as vitamin B12 levels < 20 pmol/L. 4. *, **, *** indicate significant differences (p<0.05, p<0.01, p<0.001, respectively) in the prevalence of adverse birth outcome between the micronutrient deficient or inflamed group and non-micronutrient deficient or non-inflamed group.

Adjusted RRs for maternal individual MNDs with her offspring birth outcomes are presented in **Table 2**. Deficiencies in serum folate at any time point during the pregnancy were significantly associated with increased risk of LBW (RR: 1.22, 95% CI: 1.06-1.39). Based on the data measured at the last study visit prior to delivery, serum folate deficiency (RR: 1.31, 95% CI: 1.13-1.52), vitamin A deficiency (RR: 1.21, 95% CI: 1.03-1.41) and inflammation (RR: 1.38, 95% CI: 1.13-1.68) were significantly associated with increased risk of LBW. While iron deficiency was associated with a lower risk of LBW (RR: 0.84, 95% CI: 0.72–0.98), this association was not significant after adjusting for multiple hypothesis testing. Additionally, inflammation (RR: 1.90, 95% CI: 1.27-2.85) and vitamin D deficiency (RR: 1.99, 95% CI: 1.12-3.54) were associated with higher risk of PTB.

**Table 2:** Estimated relative risks of adverse birth outcomes associated with individual micronutrient status and inflammation.

| Biomarker | Deficiency at any time point, RR(95% CI) |  |  | Deficiency at last visit, RR(95% CI) |  |  |
| --- | --- | --- | --- | --- | --- | --- |
|  | LBW | PTB | SGA | LBW | PTB | SGA |
| Inflammation | 1.11(0.95, 1.29) | 1.03(0.74, 1.44) | 1.09(0.99, 1.21) | 1.38(1.13, 1.68)** | 1.90(1.27, 2.85)** | 1.00(0.83, 1.20) |
| Iron | 0.93( 0.81, 1.07) | 1.10( 0.82, 1.46) | 0.96( 0.87, 1.06) | 0.84(0.72, 0.98) | 1.09(0.81, 1.46) | 0.93(0.84, 1.04) |
| Vitamin A | 1.17(1.00, 1.36) | 0.93(0.65, 1.34) | 1.07(0.95, 1.19) | 1.21(1.03, 1.41)* | 1.03(0.71, 1.50) | 1.05(0.93, 1.18) |
| Zinc | 1.12( 0.98, 1.27) | 1.30( 0.99, 1.71) | 1.01(0.92, 1.11) | 1.11(0.98, 1.26) | 1.35( 1.03, 1.77) | 1.02(0.93, 1.11) |
| Serum folate | 1.22(1.06, 1.39)* | 1.15(0.85, 1.56) | 1.04(0.94, 1.16) | 1.31(1.13, 1.52)*** | 1.24(0.88, 1.75) | 1.07(0.96, 1.20) |
| Vitamin D | 1.18(0.88, 1.56) | 1.32(0.72, 2.41) | 1.03(0.82, 1.29) | 0.96(0.64, 1.45) | 1.99(1.12, 3.54)* | 0.83(0.59, 1.17) |
| Vitamin B12 | 1.12(0.91, 1.38) | 1.01(0.68, 1.50) | 0.94(0.83, 1.07) | 1.09(0.91, 1.29) | 0.98(0.69, 1.38) | 0.98(0.87, 1.10) |

### Co-occurrences of MNDs and Adverse Birth Outcomes

**Supplementary Figure 4 shows in Venn diagram** the sample sizes for each individual deficiency and their overlap across five included micronutrient combinations. Figure 2 shows the percentage of LBW, PTB, and SGA stratified by different combinations of MNDs, based on any inflammation or deficiency detected at any time point during pregnancy. For the combinations of iron and vitamin A deficiency, as well as folate and vitamin B12 deficiency, women with both deficiencies had the highest prevalence of LBW and SGA compared to those with only one deficiency or none. For iron combined with vitamin B12 or folate deficiency, the highest prevalence of LBW and SGA was seen in women with only vitamin B12 or folate deficiency, respectively. Additionally, for the iron and zinc deficiency combination, the group without zinc deficiency showed higher prevalence of LBW and SGA. Overall, different MND statuses did not result in substantial differences in PTB. **Supplementary Figure 5** shows a similar pattern for the combined effects of MNDs using data from the last visit.

**Figure 2:**
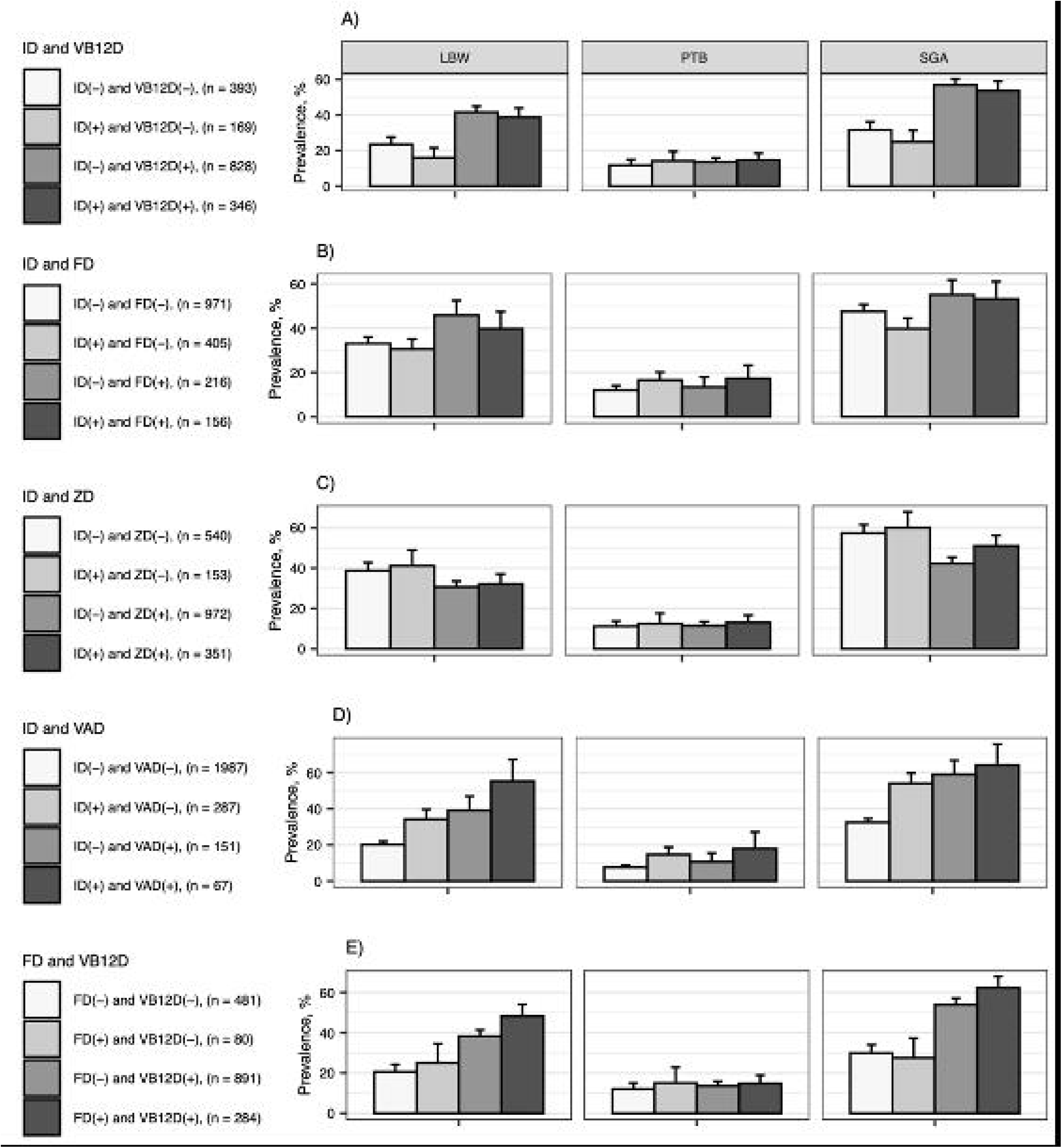
Percentages of LBW, PTB, and SGA across micronutrient deficiency combinations at any time during pregnancy: A) Iron + Vitamin B12; B) Iron + Folate; C) Iron + Zinc; D) Iron + Vitamin A; and E) Folate + Vitamin B12 1. FD, folate deficiency; ID, iron deficiency; LBW, low birth weight; PTB, preterm birth; SGA, small for gestational age; VAD, vitamin A deficiency; VB12D, vitamin B-12 deficiency; ZD, zinc deficiency. (+) means deficiency and (-) means non-deficiency. 2. LBW was defined as a birth weight of < 2500 grams; PTB was defined as a gestational age at delivery < 37 weeks; and SGA was defined as a birth weight < the 10th percentile for gestational age according to the 21st Intergrowth guidance (http://intergrowth21.ndog.ox.ac.uk/). 3. Iron deficiency was defined by inflammation-adjusted serum ferritin levels < 15 µg/L or transferrin receptor levels > 8.3 mg/L; vitamin A deficiency was defined as retinol binding protein or serum retinol levels < 0.7 µmol/L; zinc deficiency was defined based on the timing and fasting status of the blood draw, along with the zinc concentration: < 700 µg/L for fasting morning draws, < 660 µg/L for non-fasting morning draws or if fasting status was unknown, and < 590 µg/L for afternoon or unspecified times; serum folate deficiency was defined as serum folate levels < 10 nmol/L; vitamin B12 deficiency was defined as vitamin B12 levels < 20 pmol/L.

**Figure 3** displays RR estimates for LBW and SGA associated with different combinations of MNDs, reflecting inflammation or deficiency detected at any time point of pregnancy. The reference group is women without any MND (in terms of the specific combination of MND investigated). Iron deficiency but no vitamin B12 deficiency was associated with less LBW (RR: 0.40, 95% CI: 0.2-0.79). Folate deficiency without iron deficiency (RR: 1.28, 95% CI: 1.09-1.51) or without vitamin B12 deficiency (RR: 1.67, 95% CI: 1.09-2.56) was associated with an increased risk of LBW. Both folate and vitamin B12 deficiency was associated with LBW (RR: 1.38, 95% CI: 1.00-1.90). No significant associations were found for SGA or PTB. Results using data from the last study visit were consistent (**Supplementary Figure 6**) and revealed additional patterns related to iron deficiency and LBW. Specifically, women with iron deficiency in the absence of vitamin B12 deficiency (RR: 0.48, 95% CI: 0.27–0.82), folate deficiency (RR: 0.78, 95% CI: 0.65–0.95), or vitamin A deficiency (RR: 0.77, 95% CI: 0.64–0.93) had a lower risk of LBW. In contrast, women with folate deficiency in the absence of iron deficiency (RR: 1.42, 95% CI: 1.20–1.68) or vitamin B12 deficiency (RR: 1.88, 95% CI: 1.34–2.63) had a higher risk of LBW.

**Figure 3:**
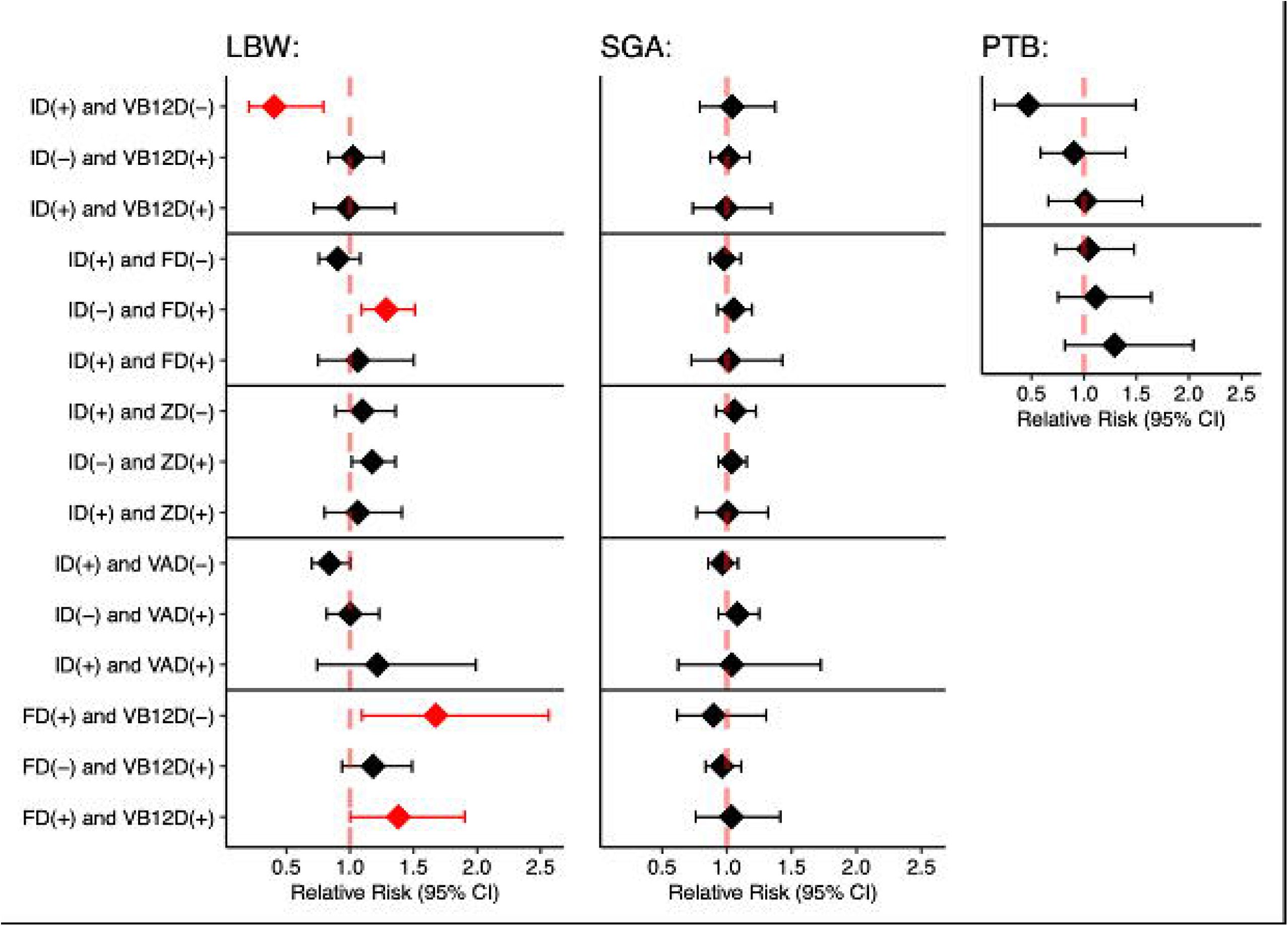
Estimated relative risks of adverse birth outcomes associated with single (in the absence of the other) or combined micronutrient deficiencies compared to women without any of the investigated deficiencies at any time point during pregnancy 1. FD, folate deficiency; ID, iron deficiency; LBW, low birth weight; PTB, preterm birth; SGA, small for gestational age; VAD, vitamin A deficiency; VB12D, vitamin B-12 deficiency, ZD, zinc deficiency; (+) means deficiency and (-) means non-deficiency. 2. LBW was defined as a birth weight of < 2500 grams; PTB was defined as a gestational age at delivery < 37 weeks; and SGA was defined as a birth weight < the 10th percentile for gestational age according to the 21st Intergrowth guidance (http://intergrowth21.ndog.ox.ac.uk/). 3. Iron deficiency was defined by inflammation-adjusted serum ferritin levels < 15 µg/L or transferrin receptor levels > 8.3 mg/L; vitamin A deficiency was defined as retinol binding protein or serum retinol levels < 0.7 µmol/L; zinc deficiency was defined based on the timing and fasting status of the blood draw, along with the zinc concentration: < 700 µg/L for fasting morning draws, < 660 µg/L for non-fasting morning draws or if fasting status was unknown, and < 590 µg/L for afternoon or unspecified times; serum folate deficiency was defined as serum folate levels < 10 nmol/L; vitamin B12 deficiency was defined as vitamin B12 levels < 20 pmol/L. 4. Analyses adjusted for age, socioeconomic status, and dataset.

## Discussion

In this large multi-country pregnancy analysis, we found a widespread micronutrient deficiencies and inflammation. Higher proportions of MNDs were observed in younger, short mothers and those with lower education level. These findings are consistent with previous research^1,27,28^ and highlight the importance of considering maternal age, height, education, and SES when addressing nutritional inequities and improving maternal health outcomes. Maternal height captures her own early childhood nutrition, genetics and transfer of intergenerational deprivation, especially among those in resource poor settings^29^. Importantly, our study explored the associations between maternal single and multiple MNDs during pregnancy and adverse birth outcomes in her offspring.

The results demonstrated that individual MNDs and inflammation are significantly associated with increased the risk of adverse birth outcomes. Among the single MNDs and inflammation, inflammation at the last study visit prior to delivery was consistently linked to higher risks of LBW and PTB. This aligns with prior studies showing that maternal inflammation is associated with fetal growth restriction and preterm labor.^30^ A counterintuitive hypothesis was observed, such that iron deficiency during late pregnancy showed a trend of protecting against LBW. Several previous studies also pointed the findings inconsistent with our hypothesis. Elevated serum ferritin level (>75th percentile, 20.2 μg/L) in late pregnancy was associated with increased risks of LBW and SGA^31^. Similarly, one study found that iron deficiency defined by serum transferrin receptor (sTfR) was paradoxically associated with a lower risk of PTB^32^. The mechanisms behind this finding remain unclear. Although most placental iron transfer from mother to fetus occurs during the third trimester^33,34^, evidence also suggests that maternal iron depletion starts as early as the second trimester^35^ – ostensibly to improve her offspring birth weight (i.e., reducing risk of LBW). Differences in plasma volume expansion, inflammation status, or other unmeasured factors may influence how micronutrients relate to birth outcomes. Vitamin A and folate deficiencies were both associated with increased risk of LBW, strengthening prior evidence on the negative impact of inadequate folate on PTB and LBW^6^. Vitamin D deficiency was associated with higher risk of PTB, in line with a meta-analysis^36^. These findings emphasize the importance of ensuring adequate maternal micronutrient levels and managing inflammation during pregnancy to reduce the risk of Adverse birth outcomes.

To our knowledge, this is the first study to evaluate how co-occurring MNDs influence adverse birth outcomes in a large multi-country dataset. Women suffering from vitamin B12 or folate deficiency had a higher prevalence of LBW (∼40%) and SGA (∼50%), regardless of iron deficiency status. Those with both vitamin B12 and folate deficiencies showed even higher prevalence of LBW (∼50%) and SGA (∼60%), suggesting a potentially additive effect. These associations were consistent across both pregnancy-wide and late-pregnancy measurements, indicating that MNDs at different gestational time points and transient nutritional demands of pregnancy may contribute to poor birth outcomes. Women with concurrent iron and vitamin A deficiencies also had higher prevalence of LBW and SGA, though only the folate and vitamin B12 combination remained significantly associated with LBW after covariate adjustment. The lack of significant associations for other MND combinations may reflect the complexity of nutrient interactions or limited statistical power due to small subgroup sizes. Folate deficiency was associated with higher LBW risk, particularly when vitamin B12 or iron status was adequate. Iron, folate and vitamin B12 all play critical roles in fetal growth and development. These perplexing findings highlight the need to further examine the underlying mechanisms to understand the role of potential nutrient interactions or other confounding factors.

A major strength of this study is the comprehensive examination of combined effects in MNDs across multiple countries and datasets. This multi-country dataset enabled us to explore variations in micronutrient status across populations and to account for pregnancy stage in our models. Furthermore, our analysis considered several combinations of MNDs, which are largely unexplored in previous research, and provide novel insights into how co-occurring MNDs over the course of influence birth outcomes. Several limitations should be acknowledged. First, micronutrient data were incomplete across datasets. Only 1 out of 11 datasets contained 6 micronutrients, while others included various subsets of micronutrient data. The limited availability of multiple micronutrient data prevented evaluation of the full spectrum of multiple MNDs comprehensively. As a result, analyses of combined deficiencies used different study subsets, limiting comparability and generalizability. Second, despite adjusting for maternal age, SES, and dataset, residual confounding may persist. Analyses accounting for additional covariates such as gestational age, and comorbidities (e.g., hypertension, diabetes) may advance the field. Third, the number of PTB cases was relatively small, possibly limiting our ability to detect significant associations. Fourth, participating trial studies varied in their nutritional supplementation interventions (e.g., iron-folic acid, multiple micronutrients, lipid-based nutrient supplements). To address this, dataset was added as a covariate to account for heterogeneity in interventions. Lastly, missing birth outcome data were present in the BRINDA pregnancy database. Approximately 38% of pregnancies (2,039 out of 5,408) lacked birth outcome information, potentially introducing bias or reducing statistical power. Of the 3,369 pregnancies with known outcomes, 2.1% were multiple births (n = 72), limiting our ability to generalize beyond singleton livebirths.

Our findings have important implications for public health policies and maternal nutrition interventions aimed at improving birth outcomes. This multi-dataset analysis revealed that maternal MNDs, particularly folate and vitamin B12 deficiencies, were associated with elevated risks of LBW, both independently and in combination. Inflammation, vitamin A and vitamin D deficiencies also contributed to adverse birth outcomes. Iron metabolism during pregnancy may be transient and, when considered alongside vitamin B12 and folate status, reflects complex nutrient interactions that may be influenced by unrecognized mechanisms. These findings highlight the need to consider multiple micronutrient pathways when evaluating maternal nutritional status and its relationship with birth outcomes.

## Data Availability

All data produced in the present study are available upon reasonable request to the authors.

## Abbreviations

ABOs: Adverse Birth Outcomes
AGP: α-1-acid glycoprotein
BRINDA: Biomarkers Reflecting Inflammation and Nutritional Determinants of Anemia
CI: Confidence Interval
CRP: C-reactive protein
DFR: False Discovery Rate
LBW: Low Birth Weight
MMN: Multiple Micronutrient
PTB: Preterm Birth
RR: Relative Risk
sTfR: Soluble transferrin receptor
SES: Socioeconomic Status
SGA: Small for Gestational Age

