## Supplementary material for "Maternal micronutrient deficiencies and inflammation and their associations with adverse birth outcomes: The BRINDA project"

Jiaxi Geng

***Supplementary Materials***

Supplementary Figure 1: Inclusion criteria and sample sizes of pregnant women

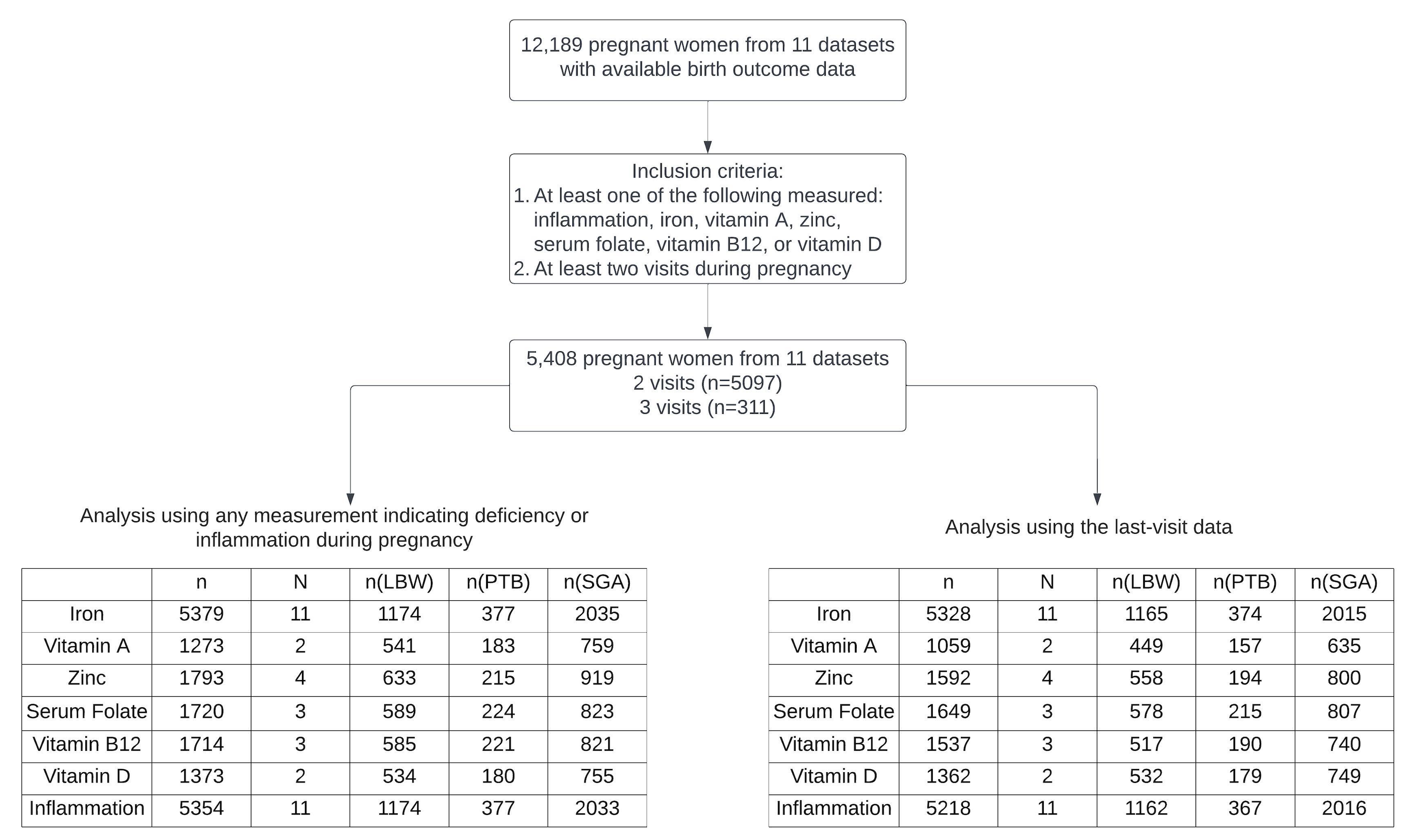

1. n denotes the sample size of pregnant women; N denotes the number of datasets.
2. LBW, low birth weight; PTB, preterm birth; SGA, small for gestational age.
3. LBW was defined as a birth weight of < 2500 grams; PTB was defined as a gestational age at delivery < 37 weeks; and SGA was defined as a birth weight < the 10th percentile for gestational age according to the 21st Intergrowth guidance (<http://intergrowth21.ndog.ox.ac.uk/>).
4. Inflammation was defined as C-reactive protein (CRP) > 5 mg/L, or α-1-acid glycoprotein (AGP) > 1 g/L; iron deficiency was defined by inflammation-adjusted serum ferritin levels < 15 µg/L or transferrin receptor levels > 8.3 mg/L; vitamin A deficiency was defined as retinol binding protein or serum retinol levels < 0.7 µmol/L; zinc deficiency was defined based on the timing and fasting status of the blood draw, along with the zinc concentration: < 700 µg/L for fasting morning draws, < 660 µg/L for non-fasting morning draws or if fasting status was unknown, and < 590 µg/L for afternoon or unspecified times; serum folate deficiency was defined as serum folate levels < 10 nmol/L; vitamin D deficiency was defined as 25-hydroxyvitamin D (25(OH)D) levels < 25 nmol/L; vitamin B12 deficiency was defined as vitamin B12 levels < 20 pmol/L.

Supplementary Figure 2: Prevalence of micronutrient deficiencies and inflammation at first visit across different demographic and risk factors

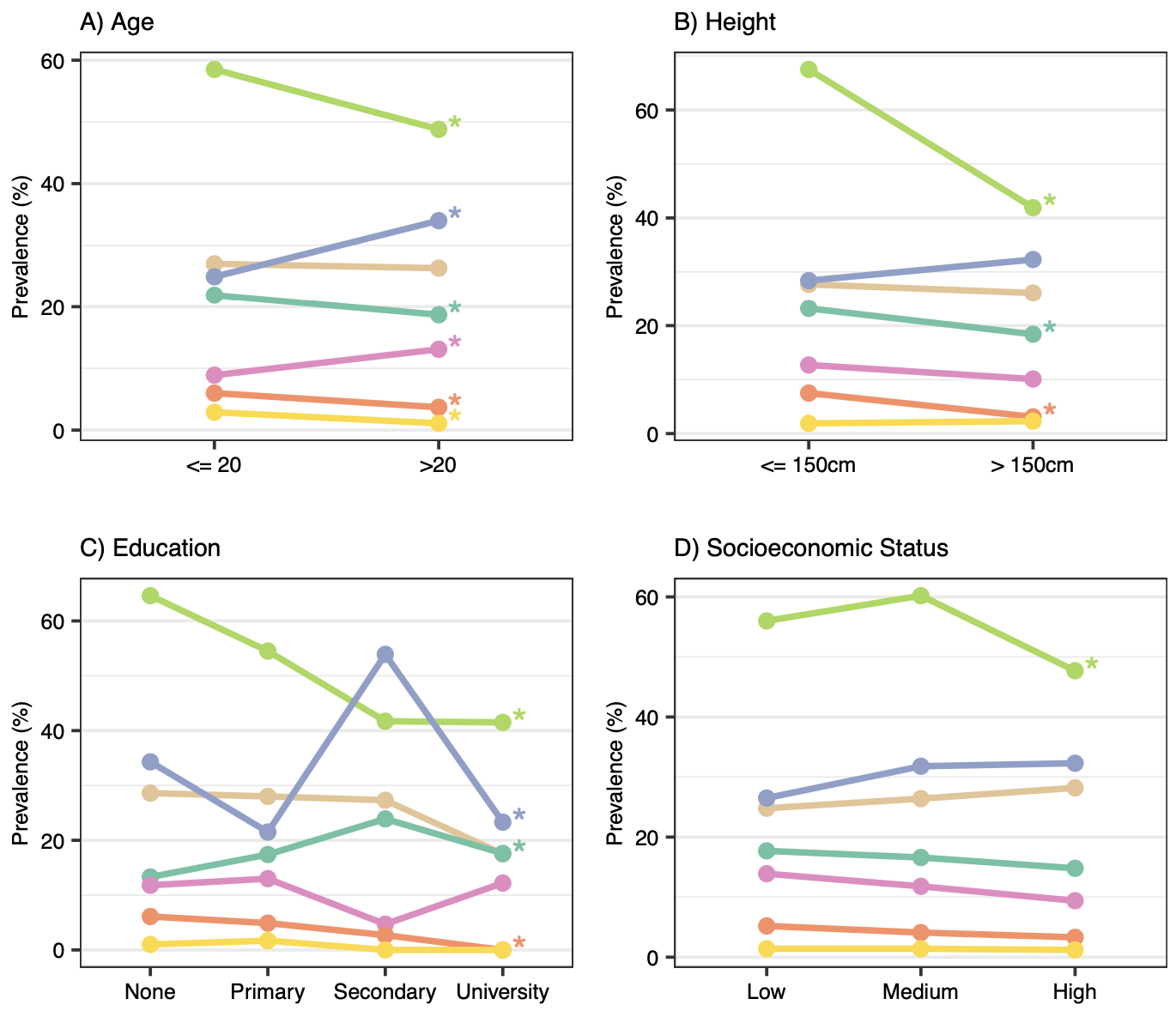

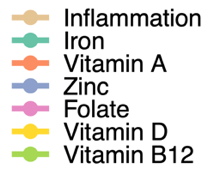

1. Inflammation was defined as C-reactive protein (CRP) > 5 mg/L, or α-1-acid glycoprotein (AGP) > 1 g/L; iron deficiency was defined by inflammation-adjusted serum ferritin levels < 15 µg/L or transferrin receptor levels > 8.3 mg/L; vitamin A deficiency was defined as retinol binding protein or serum retinol levels < 0.7 µmol/L; zinc deficiency was defined based on the timing and fasting status of the blood draw, along with the zinc concentration: < 700 µg/L for fasting morning draws, < 660 µg/L for non-fasting morning draws or if fasting status was unknown, and < 590 µg/L for afternoon or unspecified times; serum folate deficiency was defined as serum folate levels < 10 nmol/L; vitamin D deficiency was defined as 25-hydroxyvitamin D (25(OH)D) levels < 25 nmol/L; vitamin B12 deficiency was defined as vitamin B12 levels < 20 pmol/L.
2. * indicates significant associations (p<0.05) between MNDs and demographic factors using chi-square tests

Supplementary Figure 3: Percentages of three adverse birth outcomes by micronutrient deficiency and inflammation at the last visit

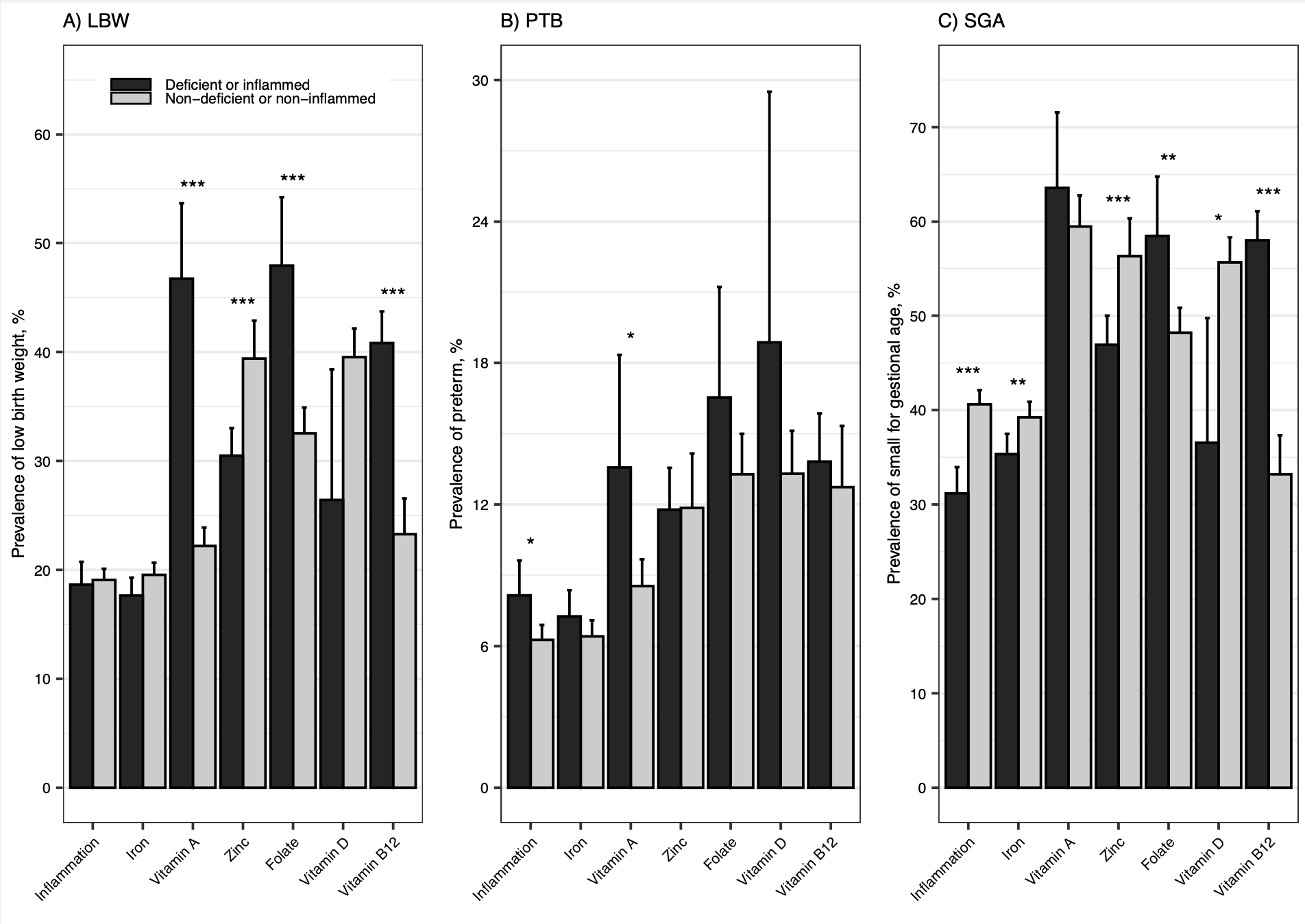

1. LBW, low birth weight; PTB, preterm birth; SGA, small for gestational age.
2. LBW was defined as a birth weight of < 2500 grams; PTB was defined as a gestational age at delivery < 37 weeks; and SGA was defined as a birth weight < the 10th percentile for gestational age according to the 21st Intergrowth guidance (<http://intergrowth21.ndog.ox.ac.uk/>).
3. Inflammation was defined as C-reactive protein (CRP) > 5 mg/L, or α-1-acid glycoprotein (AGP) > 1 g/L; iron deficiency was defined by inflammation-adjusted serum ferritin levels < 15 µg/L or transferrin receptor levels > 8.3 mg/L; vitamin A deficiency was defined as retinol binding protein or serum retinol levels < 0.7 µmol/L; zinc deficiency was defined based on the timing and fasting status of the blood draw, along with the zinc concentration: < 700 µg/L for fasting morning draws, < 660 µg/L for non-fasting morning draws or if fasting status was unknown, and < 590 µg/L for afternoon or unspecified times; serum folate deficiency was defined as serum folate levels < 10 nmol/L; vitamin D deficiency was defined as 25-hydroxyvitamin D (25(OH)D) levels < 25 nmol/L; vitamin B12 deficiency was defined as vitamin B12 levels < 20 pmol/L.
4. *, **, *** indicate significant differences (p<0.05, p<0.01, p<0.001, respectively) in the prevalence of adverse birth outcome between the micronutrient deficient or inflamed group and non-micronutrient deficient or non-inflamed group.

Supplementary Figure 4: Venn diagrams showing sample sizes for 5 included micronutrient deficiency combinations based on data indicating deficiency at any time point during pregnancy

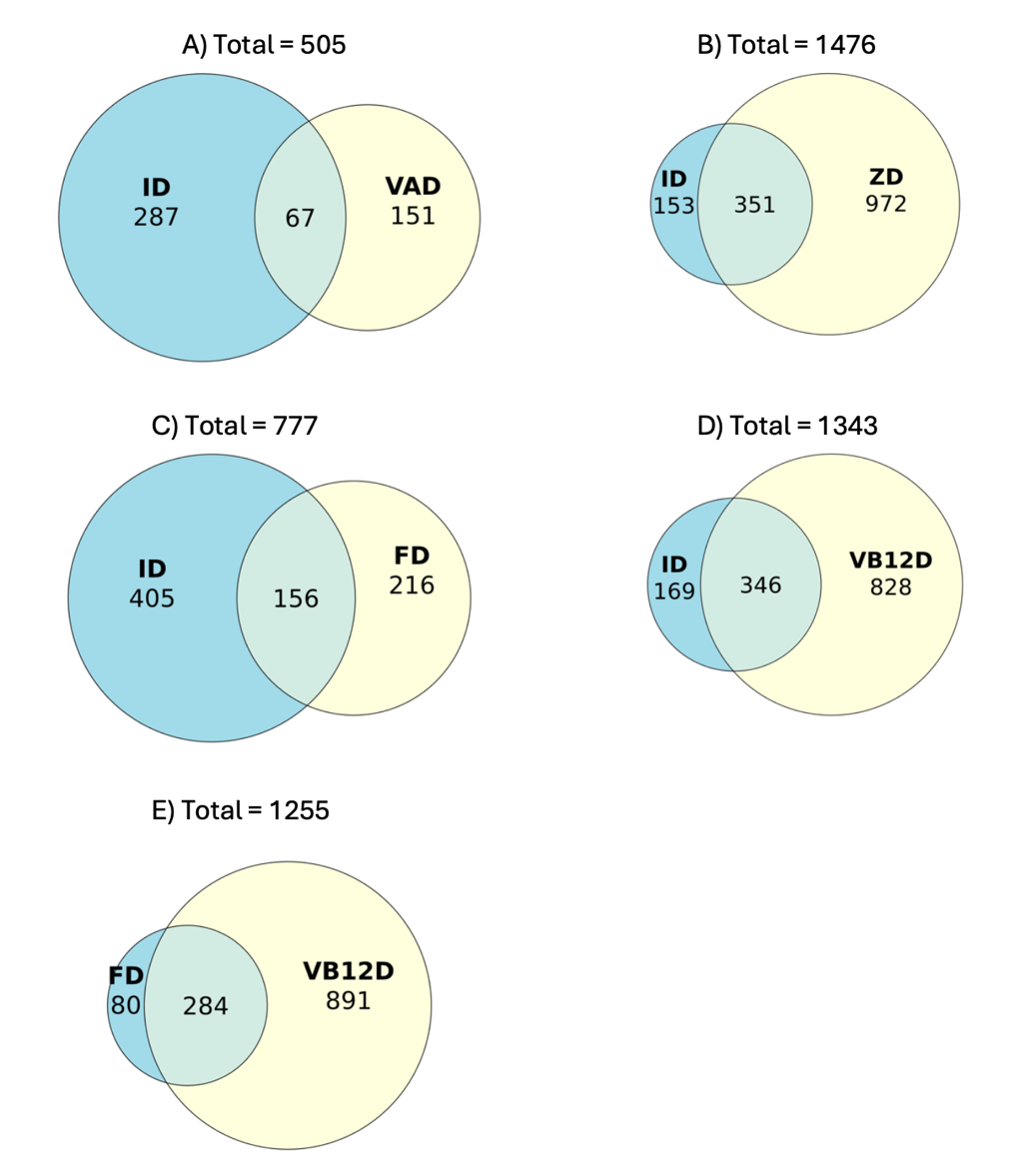

1. Figure A)-E) included data from the following datasets: A) from Bangladesh (2012) and Vietnam (2011); B) from Bangladesh (2012), Guatemala (2018), India (2017), and Pakistan (2017); C)-E) from Bangladesh (2012), Ghana (2014), and the US (2013).
2. FD, folate deficiency; ID, iron deficiency; VAD, vitamin A deficiency; VB12D, vitamin B-12 deficiency; ZD, zinc deficiency.
3. Iron deficiency was defined by inflammation-adjusted serum ferritin levels < 15 µg/L or transferrin receptor levels > 8.3 mg/L; vitamin A deficiency was defined as retinol binding protein or serum retinol levels < 0.7 µmol/L; zinc deficiency was defined based on the timing and fasting status of the blood draw, along with the zinc concentration: < 700 µg/L for fasting morning draws, < 660 µg/L for non-fasting morning draws or if fasting status was unknown, and < 590 µg/L for afternoon or unspecified times; serum folate deficiency was defined as serum folate levels < 10 nmol/L; vitamin B12 deficiency was defined as vitamin B12 levels < 20 pmol/L.

Supplementary Figure 5: Percentages of LBW, PTB, and SGA across four micronutrient deficiency combinations at the last visit: A) Iron + Vitamin B12; B) Iron + Folate; C) Iron + Zinc; D) Iron + Vitamin A; and E) Folate + Vitamin B12

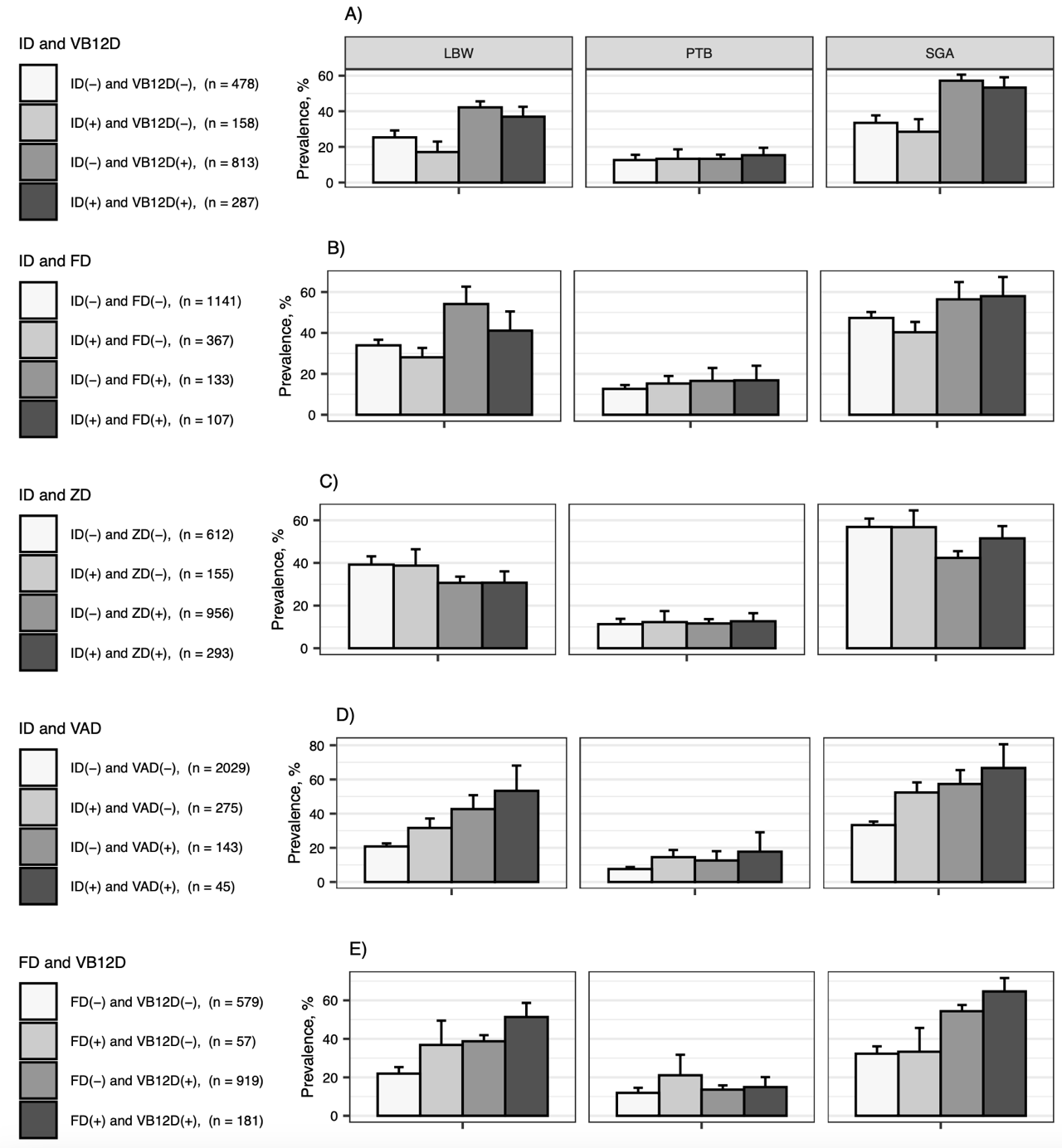

1. FD, folate deficiency; ID, iron deficiency; LBW, low birth weight; PTB, preterm birth; SGA, small for gestational age; VAD, vitamin A deficiency; VB12D, vitamin B-12 deficiency; ZD, zinc deficiency; (+) means deficiency and (-) means non-deficiency.
2. LBW was defined as a birth weight of < 2500 grams; PTB was defined as a gestational age at delivery < 37 weeks; and SGA was defined as a birth weight < the 10th percentile for gestational age according to the 21st Intergrowth guidance (<http://intergrowth21.ndog.ox.ac.uk/>).

Iron deficiency was defined by inflammation-adjusted serum ferritin levels < 15 µg/L or transferrin receptor levels > 8.3 mg/L; vitamin A deficiency was defined as retinol binding protein or serum retinol levels < 0.7 µmol/L; zinc deficiency was defined based on the timing and fasting status of the blood draw, along with the zinc concentration: < 700 µg/L for fasting morning draws, < 660 µg/L for non-fasting morning draws or if fasting status was unknown, and < 590 µg/L for afternoon or unspecified times; serum folate deficiency was defined as serum folate levels < 10 nmol/L; vitamin B12 deficiency was defined as vitamin B12 levels < 20 pmol/L.

Supplementary Figure 6: Estimated relative risks of adverse birth outcomes associated with single (in the absence of the other) or combined micronutrient deficiencies, compared to women without any of the investigated deficiencies, based on data from the last study visit

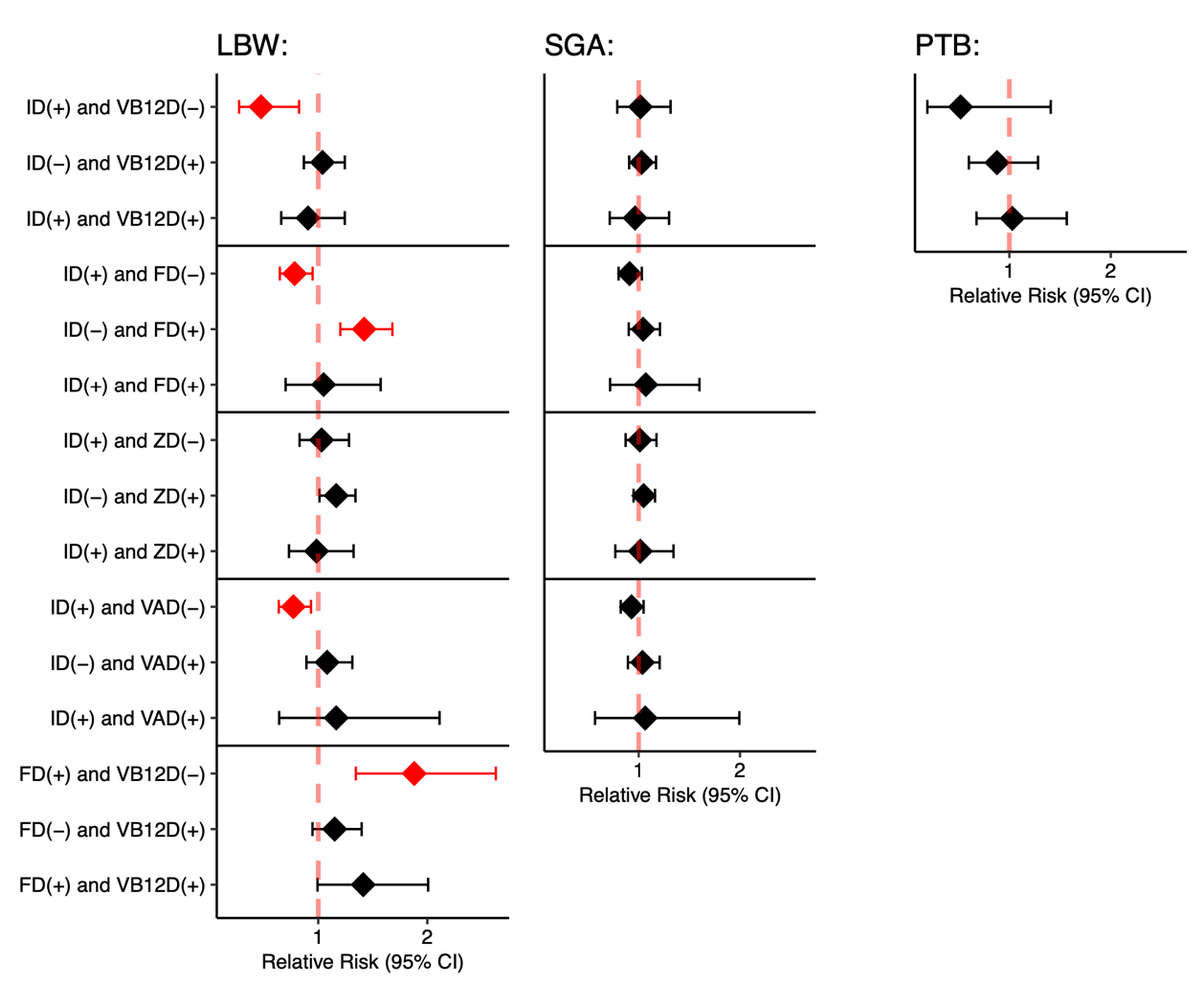

1. FD, folate deficiency; ID, iron deficiency; LBW, low birth weight; PTB, preterm birth; SGA, small for gestational age; VAD, vitamin A deficiency; VB12D, vitamin B-12 deficiency, ZD, zinc deficiency; (+) means deficiency and (-) means non-deficiency.
2. LBW was defined as a birth weight of < 2500 grams; PTB was defined as a gestational age at delivery < 37 weeks; and SGA was defined as a birth weight < the 10th percentile for gestational age according to the 21st Intergrowth guidance (<http://intergrowth21.ndog.ox.ac.uk/>).
3. Iron deficiency was defined by inflammation-adjusted serum ferritin levels < 15 µg/L or transferrin receptor levels > 8.3 mg/L; vitamin A deficiency was defined as retinol binding protein or serum retinol levels < 0.7 µmol/L; zinc deficiency was defined based on the timing and fasting status of the blood draw, along with the zinc concentration: < 700 µg/L for fasting morning draws, < 660 µg/L for non-fasting morning draws or if fasting status was unknown, and < 590 µg/L for afternoon or unspecified times; serum folate deficiency was defined as serum folate levels < 10 nmol/L; vitamin B12 deficiency was defined as vitamin B12 levels < 20 pmol/L.
4. Analyses adjusted for age, socioeconomic status, and dataset.
